# Systematic review and meta-analysis of enteric virus shedding in human excretions

**DOI:** 10.1101/2025.03.01.25323153

**Authors:** Gang Zheng, Elana M. G. Chan, Alexandria Boehm

## Abstract

**Background:** Wastewater-based epidemiology can inform the understanding of infectious disease occurrence in communities. Quantitative information on shedding of pathogen biomarkers in excretions that enter wastewater is needed to link measurements of pathogen biomarkers to rates of disease occurrence.

**Methods:** We compile, summarize, and compare data on shedding of human norovirus, rotavirus, hepatitis A, and adenovirus group F in stool, vomit, urine, saliva, mucus, and sputum using a systematic review and meta-analysis approach.

**Findings:** We provide summaries of measured concentrations of the viruses across excretions where data exist. We provide longitudinal shedding profiles in terms of concentrations and positivity rates. Duration of shedding and day of peak shedding are also provided.

**Interpretation:** There are limited data available for excretions other than stool, and limited data available for adenovirus group F. The aggregated data provided herein can serve as model inputs to translate wastewater enteric virus biomarker concentrations to disease occurrence rates. The study highlights data gaps and research needs.

## Introduction

Globally, diarrhoeal disease is a leading cause of child mortality and morbidity, causing approximately 1.7 billion cases of child diarrhea with over 400,000 deaths for children under 5 years old every year^1^. Among the most common causes of diarrhoeal diseases are enteric viruses, including norovirus, rotavirus, hepatitis A, and adenovirus group F.^2^ Despite their importance, there are limited data on the prevalence and incidence of enteric viral infections, limiting prevention efforts and response activities, and a thorough understanding of disease epidemiology.^3^

Wastewater-based epidemiology (WBE) has emerged as a tool to help better understand community health. WBE monitors pathogen biomarkers in wastewater to track infectious disease circulation in a community. Recent studies have shown enteric virus concentrations in wastewater are positively associated with traditional measures of disease occurrence.^4–6^ However, there has been limited success in translating virus biomarker concentrations in wastewater to the number of infections in a community. Mechanistic, process-based modeling studies have illustrated that information on biomarker shedding into wastewater via excreations is critical to do so.^7–10^ The goal of this study is to characterize enteric virus shedding in infected patients using a systematic review and meta-analysis approach for this application. The compiled data can be used as input parameters to models to estimate infection occurrence in communities from wastewater measurements of pathogen biomarkers, and to inform knowledge gaps and future research needs on viral shedding.

## Methods

### Search strategy and selection criteria

We reviewed the literature to report shedding of human norovirus (NoV), rotavirus (RV), hepatitis A virus (HAV), and enteric adenovirus group F (AdV) in stool, vomit, urine, saliva, mucus, and sputum. These viruses were chosen because we and others have previously measured them in wastewater and found that their concentrations there are associated with traditional disease metrics.^11–13^ The selected excretions were included as they enter domestic sewage.^14,15^ There are other excretions that may be important that we do not consider (e.g., blood, skin sloughing). The systematic review and meta-analysis were pre-registered,^16^ and followed the Preferred Reporting Items for Systematic Review and Meta-Analysis (PRISMA) guidelines.^17^

We conducted searches for peer reviewed papers using three databases using search fields as provided in Figure 1. The applied search terms included enteric virus and excretion names (appendix p 5). The exact date the search was completed is provided in appendix (p 5).

**Figure 1.**
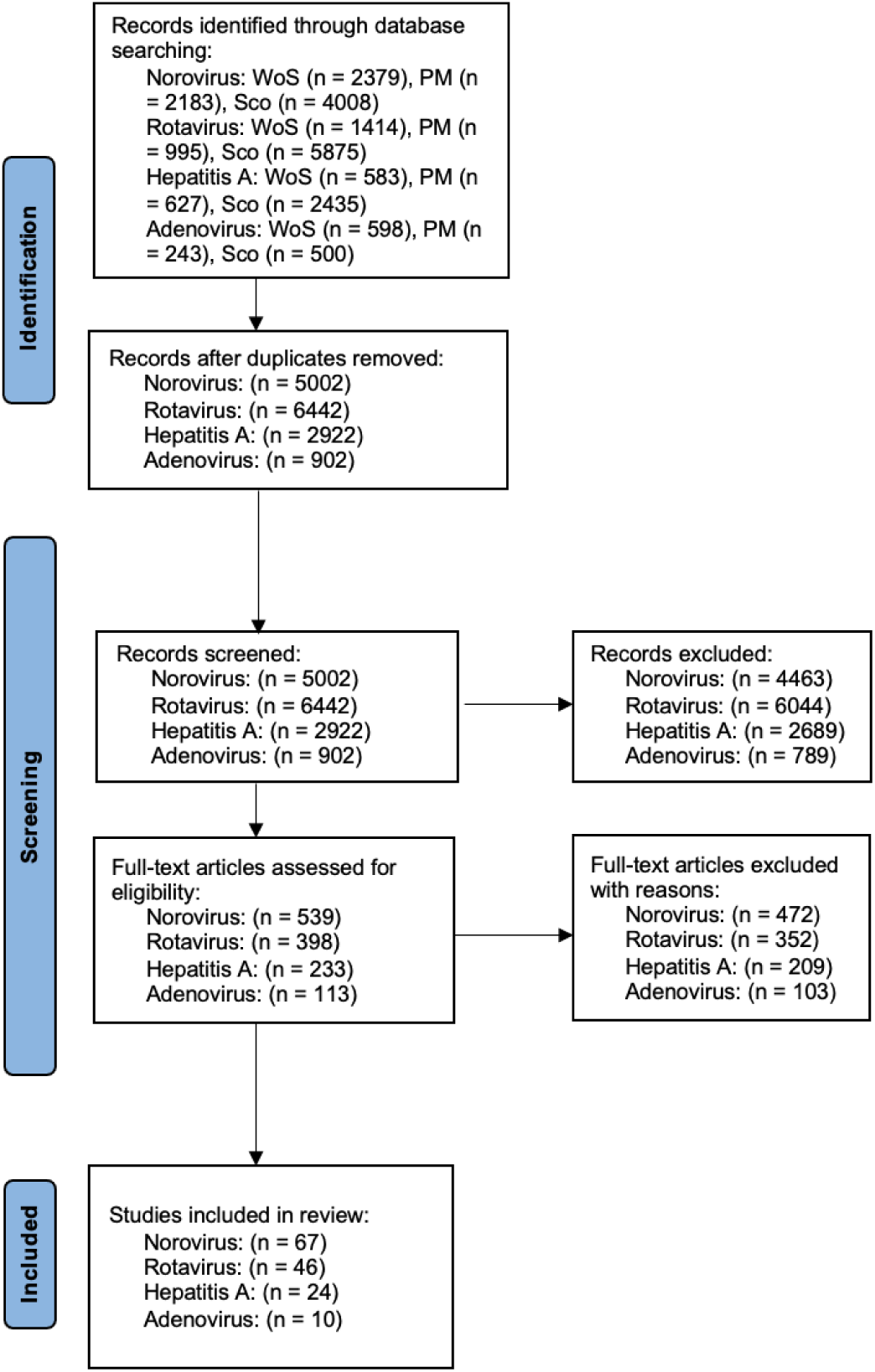
PRISMA diagram for the project.^17^ WoS is web of science, PM is PubMed, and Sco is Scopus; n is the number of papers in the categories. For each database, the following search fields were used: Web of Science (search field = All Fields), PubMed (search field = Title/Abstract), and Scopus (search field = Title/Abstract/Key).

Identified papers were imported into Covidence (https://www.covidence.org) and duplicates removed. The title and abstract of each paper were reviewed for relevance by one reviewer, and 10% of papers deemed irrelevant were reviewed independently by a second reviewer to confirm the decision making process. If papers seemed to be potentially relevant, they were advanced to full-text review. During full-text review, papers that met the following seven inclusion criteria were retained: (1) published in a peer-reviewed journal; (2) written in English; (3) contained primary data (e.g., not a review); (4) contained original data not generated by a model, estimation, or from another study; (5) contained human shedding data; (6) contained extractable shedding data, defined as concentration or longitudinal presence of the virus in an excretion; wash and swab samples were deemed valid for assessing the presence of the virus in corresponding excretions (e.g., throat swab for saliva, rectal swab for stool); and (7) contained shedding data from individuals without documented chronic infections, immunocompromised health conditions, or co-infection with multiple enteric virus. A second reviewer participated in the full-text review reviewing approximately 10% papers.

We extracted relevant information from each paper: (1) virus and excretion; (2) study type (cross-sectional or longitudinal and challenge/feeding study, natural infection, or vaccination); (3) concentrations or presence of viruses in excretion and the time at which the sample was collected (if provided), and reported units of concentration (if provided); (4) sample size; (5) virus detection method; (6) data measurement quality metrics (appendix p 6). In some papers, data were presented in the form of summary statistics without individual-level data. The type of summary statistic was recorded with all related descriptive statistics including the number of individual data used to calculate the summary statistics. For papers presenting shedding data in graphical format, WebPlotDigitizer (https://plotdigitizer.com) was used for data extraction. All extracted data are available from the Stanford Digital Repository (https://doi.org/10.25740/cs445xw7641).

Included papers were evaluated according to six quality control criteria (appendix p 6) and each assigned a quality score (QS) between 0 to 6. The paper was categorized as low confidence if QS<2, moderate confidence if 2≤QS<4, and high confidence if 4≤QS≥6. Data from papers with QS<2 were excluded.

### Data Analysis

The goals of the meta-analysis were to (1) describe observed enteric virus concentrations in excretions and (2) generate longitudinal enteric virus shedding time series in excretions both in terms of concentrations and detection positivity rates. Secondary outcomes included the duration of shedding and timing of peak shedding of viruses in each excretion.

#### Concentrations of viruses

All reported, measured concentrations of each virus in each excretion were combined, regardless of study design. Assumptions used to do so are outlined in the appendix (p 2). The resultant data were not normally distributed (Shapiro–Wilk, p<0.05), so Kruskal-Wallis tests were used to test for differences across viruses and excretions using adjusted p=0.025 to account for multiple comparisons (appendix p 2). Dunn post-hoc test with a Bonferoni correction was applied as needed.

#### Longitudinal concentration data

Longitudinal data includes the time when the excretion was obtained during the course of infection.They were categorized into two distinct groups: those reporting as a function of days after symptom (DAS) onset and days after infection (DAI). Studies reporting as a function of DAS were generally those of natural infection, while those reporting relative to DAI were generally feeding / challenge studies including vaccination studies. In compiling longitudinal data, several assumptions were made, as outlined in the appendix (p 2). Logistic models and gamma models were used to characterize shedding profiles (appendix p 7). The logistic model was defined as log_10_(conc+1)=A(1+exp(B(t-C)))^-1^ and the gamma model was log_10_(conc+1)=A t^B^exp(-Ct) where conc is concentration, t is time, and A, B, and C are fitting parameters. Models were fit using nonlinear least squares (NLS) optimization via the nlsLM function from the minpack.lm package in R (4.4.0). Longitudinal concentration datasets with smaller than 5 discrete days of data were excluded from plotting and shedding profile modelling, as we deemed this number of data points insufficient.

The timing of peak viral shedding was defined as the time at which the highest viral concentration occurred during the shedding process. Peak viral shedding day was recorded for each minimal data category provided within the paper (i.e., an individual patient time series or an aggregated time series). Aggregated peak shedding days were not normally distributed (Shapiro–Wilk, p<0.05), so we used Kruskal-Wallis to compare across viruses and excretions using p=0.05 as only one set of comparisons was possible due to data limitations.

#### Longitudinal presence data

Longitudinal presence data are observations regarding the presence or absence of viruses in excretions over time. Longitudinal concentration studies were incorporated into the longitudinal presence data. Longitudinal presence data were categorized as relative to DAS or DAI, with several additional assumptions in the appendix (p 2) regardless of study design.

We described virus presence at each available time point using positivity rates: 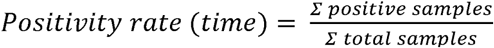. For time points where the number of data points was less than 5, the positivity rate was not calculated, as we deemed that such a small sample size would provide a positivity rate with too much uncertainty. Logistic models and gamma models were used to characterize shedding profiles. The logistic and gamma models are defined as above, except on the left hand side of the equation is positivity rate. Datasets with smaller than 5 discrete days of data were excluded for plotting and shedding profile modelling.

The duration of viral shedding, defined as the time interval between day 0 to the last positive sample, was determined from longitudinal presence data. The aggregated duration variable was not normal (Shapiro–Wilk, p<0.05), so Kruskal-Wallis test was used to evaluate differences in the shedding duration across different viruses, with a p value adjusted for multiple comparisons (appendix pp 2-3). The Dunn post-hoc test with a Bonferoni correction was also used.

To address the potential for publication bias toward selective reporting of results, we used the proportion of reported positive results in each included paper as an indicator of potential bias:

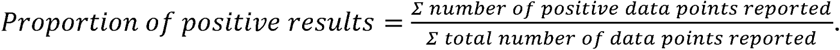

A proportion between 0.33 to 0.66 would indicate no selective reporting, as studies consistently report both negative and positive results. Additional details are in the appendix (p 3).

The funders of the study had no role in study design, data collection, data analysis, data interpretation, or the writing of the report. The corresponding author had full access to all data in the study and final responsibility for the decision to submit this systematic review and meta analysis for publication.

## Results

### Systematic review

A total of 5002, 6442, 2922 and 902 papers for NoV, RV, HAV, and AdV respectively, were identified from the three databases after dereplication. Following the screening process, 67 papers on NoV, 46 on RV, 24 on HAV, and 10 on AdV (147 papers total) qualified for data extraction (Figure 1). Out of the 147 papers, 73% (n = 108) reported sampling methodologies, 100% (n = 147) reported sample detection methods, 61% (n = 89) reported a detection threshold, 27% (n = 40) reported use of controls, 22% (n = 33) reported use of replications, and 54% (n = 80) reported sample preservation and processing (appendix p 8). Four papers (1 NoV, 2 RV, 1 HAV) were categorized as low confidence and excluded leaving 153 included papers (appendix p 3).

The total number of data points for each virus and excretion varied between 0 and 14186 (appendix p 9). The largest number of data points were available for stool (n = 26642), with comparatively fewer data points (n < 200) available for other excretions. A larger number of data points were available for NoV (n = 7938) and RV (n = 16496) compared to HAV (n = 2334) and AdV (n = 227). A summary of the study types and measurement approaches used are in the appendix (p 3).

### Norovirus

NoV concentrations were reported in stool (44 papers),^18–61^ vomit (2 papers),^55,62^ and saliva (1 paper).^63^ Median NoV concentration in these excretions was 7.7 log_10_ copies per gram stool (interquartile range (IQR) = 6.1-9.0; n = 2368), 4.6 log_10_ copies per milliliter vomit (IQR = 4.5–5.8; n = 104), and 3.3 log_10_ copies per milliliter saliva (IQR = 2.9-3.5; n = 7) (Figure 2). No data were found on NoV concentration in urine, mucus, and sputum.

**Figure 2.**
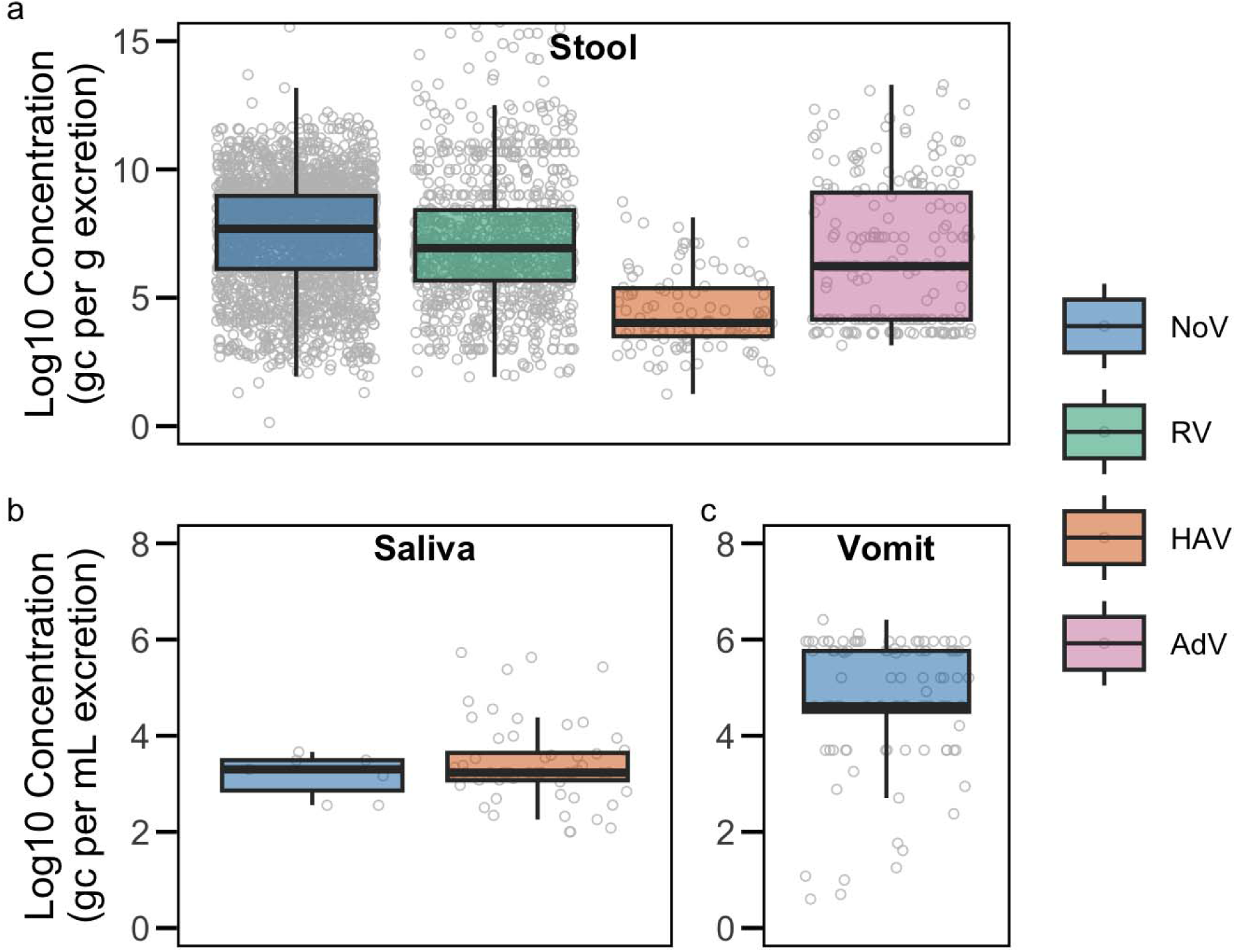
Concentration of various enteric viruses in different excretions: a) stool, b) saliva, and c) vomit. Individual concentration data points are represented as gray circles. Data points exceeding 15 loglJlJ genome copies (gc) per g/mL excretion are not shown in this figure. The lower and upper edges of the boxes correspond to the 25th and 75th percentiles, respectively, while the center line represents the median. Whiskers extend to the smallest and largest values within 1.5 times the IQR. Abbreviations: NoV = norovirus, RV = rotavirus, HAV = hepatitis A virus, AdV = adenovirus.

Longitudinal NoV concentrations were measured in stool relative to DAS (11 papers)^22,23,29,33,36,39,46,49,51,60,64^ and relative to DAI (3 papers),^28,45,55^ in saliva relative to DAS (1 paper),^63^ and in vomit relative to DAI (1 paper).^55^ No longitudinal concentration data were available for other excretions. We applied statistical models to the profiles for stool and saliva with respect to DAS and DAI (Figure 3, appendix pp 10-11). Vomit was not modeled or plotted due to the limited number of discrete days (< 5 discrete days). Median peak NoV shedding in stool occurred 3.0 DAS (IQR = 1.0-5.0; n = 40) and 3.7 DAI (IQR = 3.0-4.7; n = 44). The median peak NoV shedding day was 1.0 DAI (IQR = 1.0-1.3; n = 4) in vomit and 6.0 DAS (n = 1) in saliva (Figure 4).

**Figure 3.**
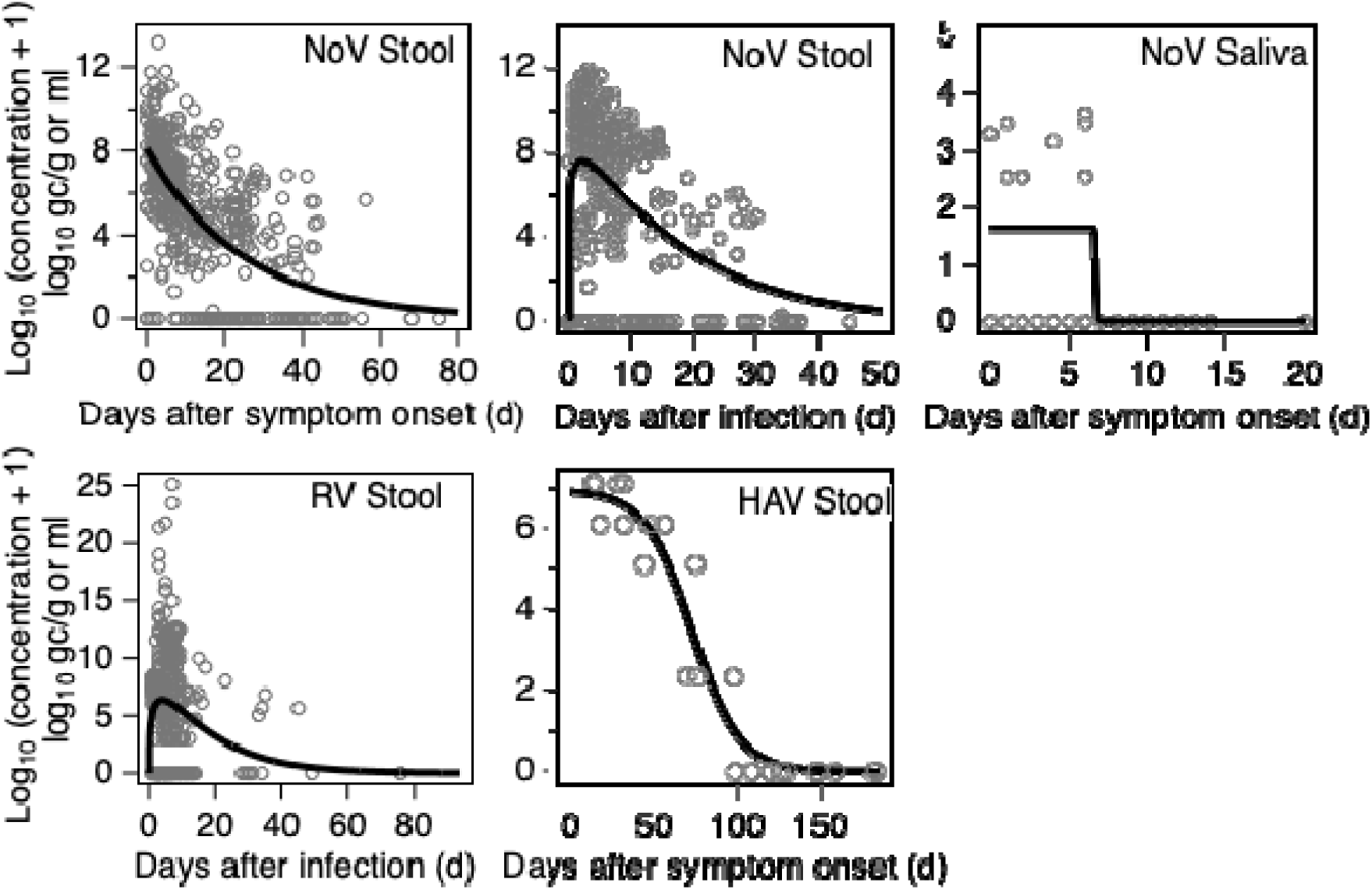
Longitudinal concentration profiles of available data on enteric virus shedding in excretions. Only data on NoV, RV, and HAV are available for stool, and only NoV is available in saliva. No other viruses or excreations had sufficient data for plotting. Profiles are plotted as a function of days after symptom onset or days after infection, depending on data availability. Concentrations are expressed log_10_ gene copies (gc) per gram (stool) or ml (saliva); all values were measured by quantitative PCR methods. Grey circles are all raw data extracted from papers on concentrations as a function of time. Black line is the modeled concentration profile. Model parameters are in appendix pp (10-11). Abbreviations: NoV = norovirus, RV = rotavirus, HAV = hepatitis A virus, AdV = adenovirus.

**Figure 4.**
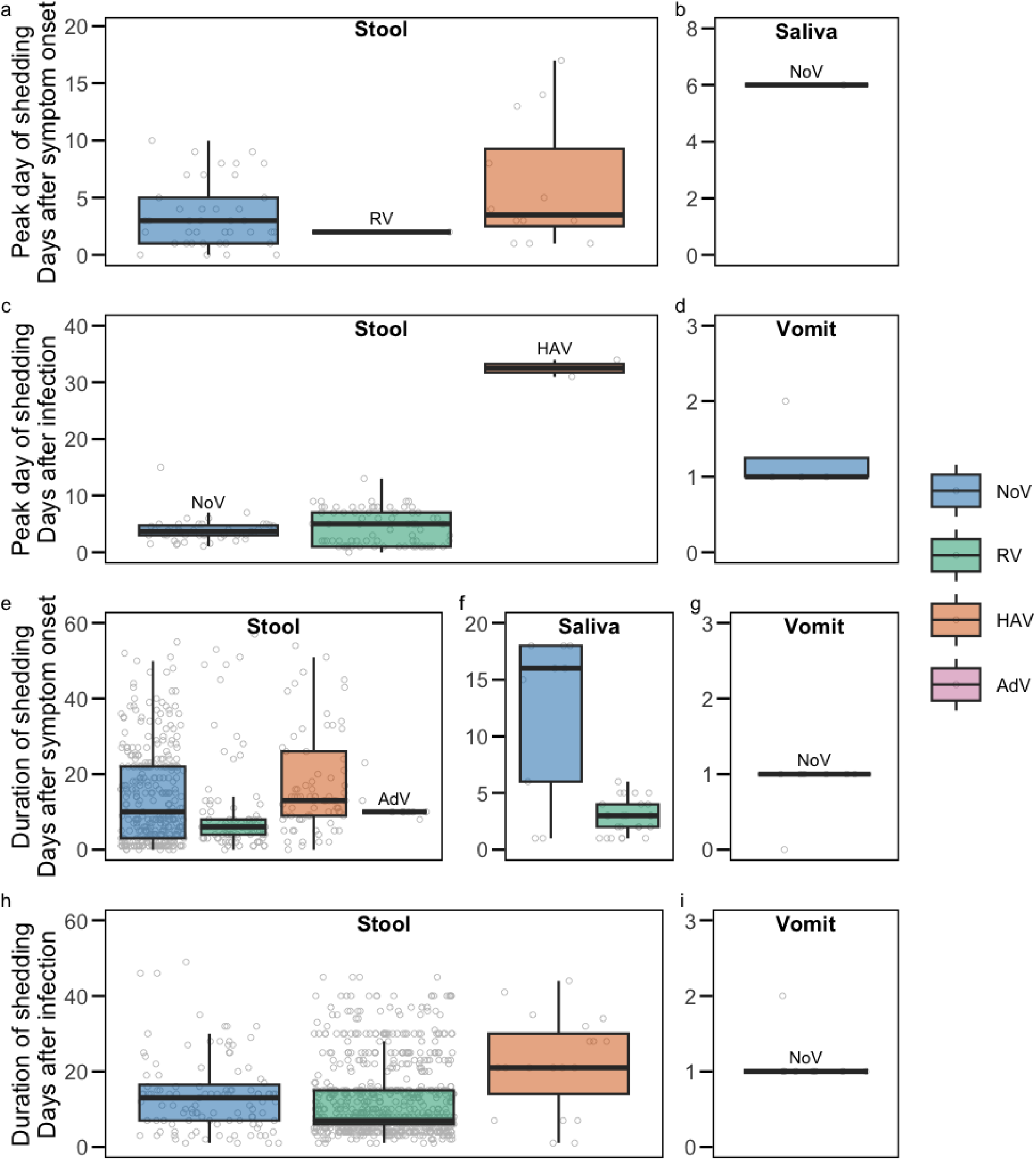
Peak day of shedding and duration of shedding in units of days for different enteric viruses in (a-d) Peak day of shedding in stool and vomit, measured relative to days after symptom onset or infection. (e-i) Duration of shedding in stool, saliva, and vomit, relative to days after symptom onset or infection. Gray circles represent individual data points. The lower and upper edges of the boxes correspond to the 25th and 75th percentiles, respectively, while the center line represents the median. Whiskers extend to the smallest and largest values within 1.5 times the IQR. Abbreviations: NoV = norovirus, RV = rotavirus, HAV = hepatitis A virus, AdV = adenovirus.

Longitudinal NoV presence data was provided in stool relative to DAS (25 papers)^18,22,23,29,33,36,39,46,49,51,60,65–78^ and DAI (10 papers),^28,39,45,55,79–84^ in saliva relative to DAS (3 papers),^63,77,85^ and in vomit relative to DAS (3 papers)^66,75,77^ and DAI (2 papers).^55,86^ We modeled the profiles for stool and saliva (Figure 5, appendix p. 10-11). Vomit was not modeled or plotted due to insufficient data (< 5 discrete days). Median NoV shedding duration in stool was 10.0 DAS (IQR = 3.0-22.0; n = 320) and 13.0 DAI (IQR = 7.0-16.5; n = 107). Median NoV shedding duration in vomit was 1.0 DAS (n = 7) and 1.0 DAI (n = 8). Median shedding duration for NoV in saliva was 16.0 DAS (IQR = 6.0-18.0; n = 9) (Figure 4). No studies provided longitudinal presence data on NoV in urine, mucus, or sputum.

**Figure 5.**
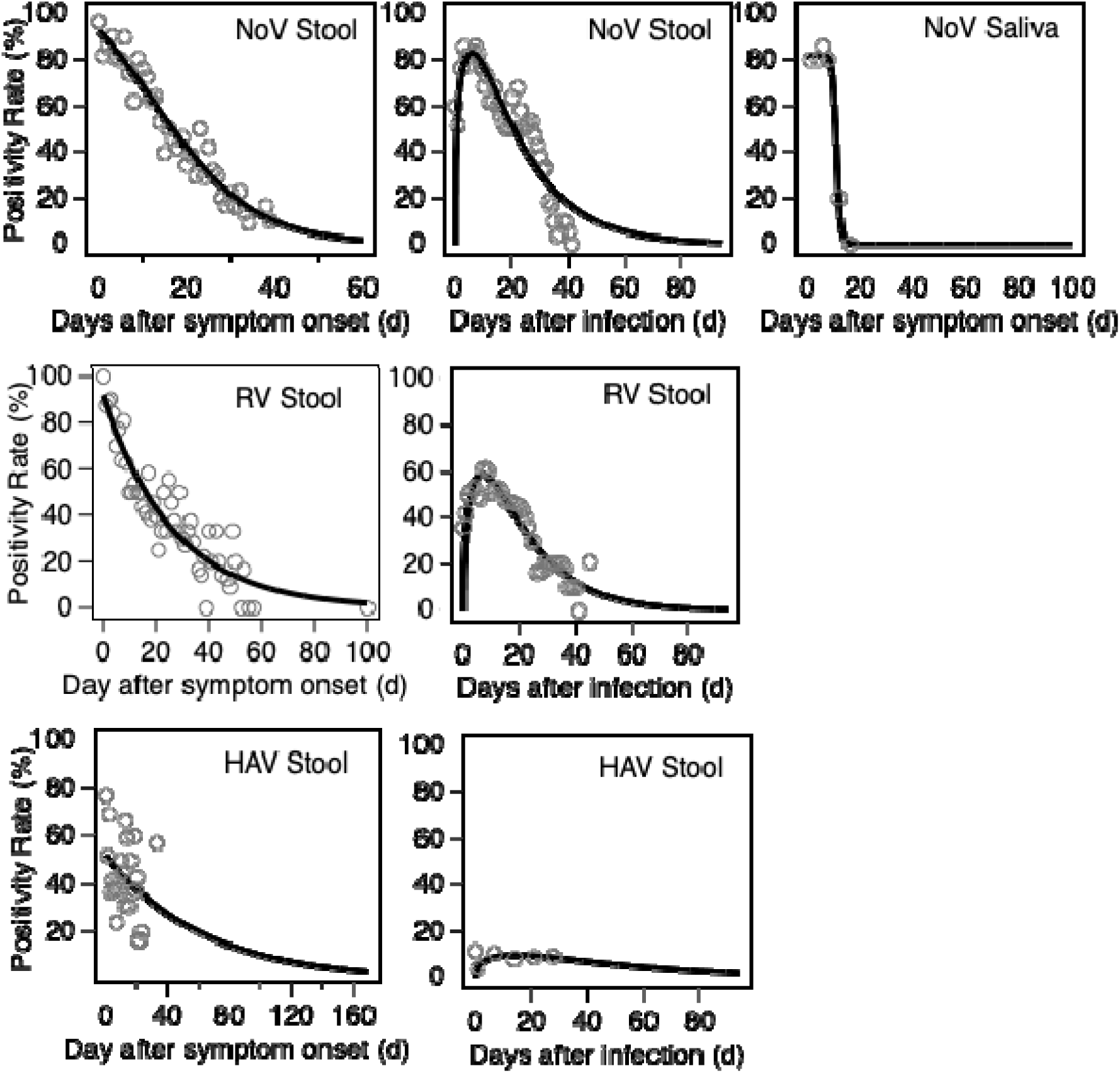
Longitudinal positivity rate profiles of available data on enteric virus shedding in excretions. Only data on NoV, RV, and HAV are available for stool, and only NoV is available in saliva. No other viruses or excreations had sufficient data for plotting. Profiles are plotted as a function of days after symptom onset or days after infection, depending on data availability. Grey circles represent combined positivity rates calculated for each day as a function of time. Black line is the modeled positivity rate profile. Model parameters are in appendix pp 10-11. Abbreviations: NoV = norovirus, RV = rotavirus, HAV = hepatitis A virus, AdV = adenovirus.

#### Rotavirus

Fifteen papers presented data on RV concentration in stool.^35,46,50,56,87–97^ Median RV concentration in stool was 6.9 log_10_ copies per gram (IQR = 5.7-8.4; n = 988) (Figure 2). No paper reported concentration data in other excretions.

Longitudinal RV concentration data in stool was measured as a function of DAS (1 paper)^46^ and DAI (5 papers).^87,89,90,93,96^ Profiles and their models (appendix pp 10-11) are provided in Figure 3. Median peak day of RV shedding in stool occurred 2.0 DAS (n = 1) and 5.0 DAI (IQR = 1.0-7.0; n = 75) (Figure 4).

Longitudinal RV presence data was provided in stool as a function of DAS (12 papers)^46,91,98–107^ and DAI (22 papers)^87,90,93,96,108–125^ and in saliva as a function of DAS (2 papers).^91,103^ Statistical models were applied to the stool profiles (Figure 5, appendix pp 10-11). Saliva data were not modeled or plotted because data were only available for a limited number (< 5) days. Median RV shedding duration in stool was 6.0 DAS (IQR = 4.0-8.0; n = 107) and 7 DAI (IQR = 6.0-15.0; n = 833). Median RV shedding duration in saliva was 3.0 DAS (IQR = 2.0-4.0) (n = 25) (Figure 4). While none of the studies investigated RV shedding in excretions beyond stool and saliva, a limited number of reports (which did not fit our inclusion criteria) noted the detection of RV RNA in other excretions, such as vomit,^126,127^ urine,^128,129^ and nasal secretions.^130–132^

#### Hepatitis A

HAV concentration data were provided in stool (4 papers)^133–136^ and saliva (4 papers).^137–140^ Median HAV concentration was 4.0 log_10_ copies per gram stool (IQR = 3.5-5.4; n = 104) and 3.2 log_10_ copies per milliliter saliva (IQR = 2.9-3.5; n = 56) (Figure 2). No data were identified regarding HAV concentration in other excretions.

One paper reported longitudinal concentration data for HAV shedding in stool measured as a function of DAS and the profile was modeled (Figure 3, appendix pp 10-11).^134^ Median peak day of HAV shedding in stool was 3.5 DAS (IQR = 2.5–9.3; n = 12) and 32.5 DAI (n = 2) (Figure 4).

Longitudinal HAV presence data for stool was provided as a function of DAS (12 papers)^134,141–151^ and DAI (4 papers).^152–155^ These data were modeled (Figure 5 and appendix pp 10-11). Median HAV shedding duration in stool was 13.5 DAS (IQR = 9.0-26.0; n = 81) and 21.0 DAI (IQR = 14.0-30.0; n = 19) (Figure 4). Although no studies included in the analysis reported HAV shedding in excretions other than stool and saliva, one study (not meeting our inclusion criteria) documented HAV RNA presence in urine.^156^

#### Adenovirus Group F

Ten papers provided concentration data on AdV in stool.^35,50,88,157–163^ Median AdV concentration in stool was 6.2 log_10_ copies per gram (IQR = 4.2–9.1; n = 214) (Figure 2). Concentration data were not available for other excretions.

One paper reported longitudinal AdV concentration data in stool as a function of DAS.^157^ For longitudinal presence data, 2 studies provided data for stool as a function of DAS.^157,164^ As both the longitudinal concentration and presence data had a limited number of discrete days (< 5 discrete days), they were not plotted or modeled. Median AdV shedding duration in stool was 10.0 DAS (IQR = 10–10; n = 10) (Figure 4). While no studies investigated AdV shedding in excretions other than stool, there were reports (not meeting our inclusion criteria) documenting the AdV presence in saliva,^165^ and urine.^166,167^

### Meta-analysis

We compared concentration across excretion types for each virus. Median NoV concentration was highest in stool followed by vomit and saliva (Figure 2). NoV concentrations were significantly different across excretion types (Kruskal-Wallis test, p<0.0001). Post hoc tests indicated significant differences in concentrations between stool and saliva, and stool and vomit (both p<0.0001). Median HAV concentration was significantly higher in stool compared to saliva (Figure 2, p<0.0001). No statistical test was conducted to compare virus concentrations of RV and AdV across different excretions as concentration data for these viruses were only available for stool.

We compared concentrations across viruses for each excretion type. In stool, median NoV concentration was highest, followed by RV, AdV, and HAV (Figure 2); concentrations were different (p<0.0001). Post hoc analysis indicated statistically significant differences between NoV and HAV, NoV and AdV, AdV and HAV, HAV and RV, and NoV and RV (all p<0.0001). There were no differences in concentrations of NoV and HAV measured in saliva (p=0.074). Statistical comparisons for other excretions were not conducted due to lack of data.

We compared peak day across excretions for each virus. For data reported relative to DAI, the peak shedding day of NoV occurred 2.7 days earlier in vomit than in stool, and the difference in peak day was statistically significant (p=0.0025). It was not possible to compare the peak shedding day for other viruses due to insufficient (n < 2) or lack of data.

We compared peak day across viruses for each excretion. For data reported relative to DAS, sufficient data (n > 2) for NoV and HAV allowed for a comparison; there was no difference. For data reported relative to DAI, peak shedding day in stool occurred earliest for NoV (3.7 days), followed by RV (5.0 days) and HAV (32.5 days); differences were statistically significant (p=0.044). Post hoc analysis indicated statistically significant differences between NoV and HAV (p=0.019) and RV and HAV (p=0.027). Data was unavailable for other comparisons.

We compared shedding duration across excretions for each virus. For data reported as a function of DAS, the duration of shedding for NoV was the longest in saliva (16 days), followed by stool (10 days) and vomit (1 day) (Figure 4), and the durations were statistically different (p=0.00075). Post hoc tests indicated significant differences in shedding duration between stool and vomit (p=0.00023) and saliva and vomit (p=0.011). For data reported relative to DAI, the shedding duration for NoV in stool (13 days) was significantly longer compared to vomit (1 day) (p<0.0001). For RV, shedding duration relative to DAI was significantly longer in stool (6 days) compared to saliva (3 days) (p<0.0001). Other comparisons were not possible due to lack of data.

We compared shedding duration across viruses for each excretion. The duration of shedding in stool relative to DAS was longest for HAV (13.5 days) followed by NoV (10.0 days), AdV (10 days), and RV (6 days) (Figure 4); durations were significantly different (p<0.0001). Post hoc tests indicated significant differences in shedding duration in stool between NoV and HAV (p=0.0035), RV and HAV (p<0.0001), and RV and NoV (p=0.0017). For saliva, shedding duration relative to DAS was significantly longer for NoV (16 days) than RV (5 days) (p=0.0072).

For data reported relative to DAI, shedding duration in stool was longest for HAV (21 days) followed by NoV (13 days) and RV (7 days) (Figure 4); the durations were significantly different (p<0.0001). Post hoc tests showed statistically significant differences between NoV and RV (p= 0.012) and between RV and HAV (p=0.00065). Other comparisons were not possible owing to lack of data.

### Publication Bias

We found no evidence of publication bias. Overall median publication proportion of positive results was 0.57 (IQR = 0.35-0.79, n = 143), indicating no strong bias toward selectively reporting either positive or negative outcomes. Additional information is in the appendix (p 4).

## Discussion

There are limited cross sectional and longitudinal data on enteric virus shedding in excretions we considered in this study. AdV data were most limited while NoV data were most available. Stool concentrations were available for each virus while we found very limited to no concentrations in saliva, sputum, urine, mucus, and vomit. Additional research is needed to measure viral concentrations in these excretions to inform interpretation of their concentrations in wastewater for WBE applications.

Only HAV and NoV concentrations in excretions other than stool were available in the literature. For NoV, concentrations tended to be highest in stool compared to other excretions (vomit and saliva). On a per mass basis, stool tended to have 1000 times higher NoV concentrations than other excretions. In contrast, HAV concentrations in stool and saliva tended to be similar. The flux of stool (mass per day) into wastewater is expected to be higher than that of vomitus or saliva on an individual basis which might suggest that of these three excretions,^168^ stool is the main excretion contributing NoV and HAV to wastewater. A mechanistic modeling study, like that of Chen and Bibby,^168^ could assess the relative importance of different excretions in contribution biomarkers to wastewater; however, such an analysis is beyond the scope of the current study.

We previously conducted a review of respiratory virus shedding in excretions that contribute to wastewater.^169^ In that study, we also identified a lack of quantitative data, with the most available for influenza. In contrast to the findings here for NoV, where stool had higher concentrations than saliva by three orders of magnitude, influenza concentrations were higher in saliva compared to stool by two to three orders of magnitude. This is perhaps not unexpected as NoV infects the gastrointestinal tract and influenza primarily the respiratory tract. Interestingly, concentrations of NoV in wastewater tend to be much higher than concentrations of influenza.^6,170^

Median shedding durations varied between 1 and 16 DAS or 1 and 21 DAI depending on virus and excretion. HAV tended to have the longest shedding duration (13 DAS or 21 DAI) among all the viruses. Shedding duration defines the length of time an infected individual may contribute biomarkers to wastewater via a specific excretion and therefore the duration that a biomarker of disease may be detected in wastewater. For example, the long duration of shedding of HAV may suggest that detection of HAV in wastewater could indicate the presence of convalescing individuals in a sewershed rather than acutely ill individuals.

In our study, we chose to not combine data measured as a function of DAS and DAI as the time period between infection and symptom onset (incubation time) can be highly uncertain and vary among individuals.^171^ By subtracting median shedding duration for the same virus and excretion measured as a function of DAS and DAI, we can estimate incubation time. This was only possible using stool for NoV, HAV, and RV, and we calculated incubation time to be 3, 8, and 1 days, respectively. These values generally agree with studies that state incubation times for RV and NoV are short (less than 48 hours),^172,173^ while incubation time for HAV is long (15-50 days).^174^ In our review of the literature, we also found longitudinal data measured as a function of days after detection. Given the uncertainties associated with interpreting this measure of time, we omitted such data from our analysis.

The longitudinal data provided in this paper can be used to model the prevalence, reproductive numbers, and incidence forecasts of different diseases.^175–177^ Similarly, concentration data can be used to inform estimates of the number of infected individuals contributing biomarkers to wastewater.^8,178–180^ Aside from using these data to improve interpretation of biomarker concentrations in wastewater, the concentration data summarized herein may also be useful for microbial risk assessments where hazard identification and characterization is the first step.^181,182^

It is important to note that many of the papers we included in this study were lacking in one or more quality control metrics. In particular, many studies omitted data on laboratory methods including the use of positive and negative controls and replication and study detection limits. Such information is essential when reporting on measurements of viruses and we recommend that future work in this area use accepted data reporting guidelines such as the Minimum Information for Publication of Quantitative Real-Time PCR Experiments (MIQE) or the Environmental Microbiology Minimum Information (EMMI) guidelines for PCR measurements.^183,184^ We found that some studies reported pseudo virus concentrations by reporting Cq values from quantitative PCR machines. Without detailed information on standard curves and other methodological details, these values have limited external validity and therefore could not be used to inform our analysis of concentrations in excretions. We urge laboratories to report quantitative data according to the MIQE or EMMI guidelines so that they are externally valid.^183,184^

There are several limitations associated with this work. First, it is important to note that there were different and limited data available across viruses and excretions which may lead to biases in our analyses that compare such measurements. Some studies reported paired data, and we did not omit those from our meta-analysis, doing so yielded the same results as including them (appendix p 4). Second, we did not consider the role of dose in affecting shedding in studies that were viral feeding and challenge studies. Third, we combined data from studies conducted using different measurement methods for the analyses. Fourth, we did not differentiate between studies conducted using vaccines; for example, much of the RV longitudinal studies focused on post-vaccination shedding in stool. Finally, the data reported herein likely under-represent shedding from asymptomatic natural infections, which would not be captured by the study designs implemented by researchers. We recommend future studies that target this population. All data available from this review are available for other researchers should they desire to conduct further analyses to account for some of these limitations.

## Research in context

### Evidence before this study

Wastewater-based epidemiology is a valuable tool for monitoring infectious diseases at the community level. Previous research has demonstrated the presence of enteric viruses in wastewater and shown a link between measurements of pathogen biomarkers to rates of disease occurrence. However, the interpretation of wastewater viral concentrations for public health applications remains challenging due to the lack of well-characterized viral shedding profiles. To address this gap, we conducted a comprehensive literature review research on Web of Science, PubMed, and Scopus with defined keywords to identify studies reporting viral shedding data in stool, vomit, saliva, urine, mucus, and sputum for norovirus, rotavirus, hepatitis A, and adenovirus group F. Studies meeting inclusion criteria were evaluated for methodological data quality and risk of selective reporting bias, and subsequently, data were extracted. Our search identified over 100 studies reporting viral concentrations, and longitudinal concentration and presence/absence data across different viruses and excretions. No prior studies have systematically compiled enteric virus shedding data for these viruses and excretions nor provided quantitatively modeled their shedding profiles.

### Added value of this study

This study is the first to systematically characterize enteric virus shedding in infected patients using a systematic review and meta-analysis approach. We identified variations in the shedding concentrations, day of peak shedding, and shedding duration between different excretions and viruses. This study provides statistical models of shedding profiles to describe the temporal variation in enteric viruses positivity/concentration in excretions. The information provided in the study can be used in mass-balance, physical models to assess disease incidence from wastewater concentrations of the viruses.

### Implications of all the available evidence

This review and meta-analysis provides quantitative shedding profiles and key parameters for modeling infection occurrence from wastewater measurements. Our findings highlight substantial gaps in the literature, particularly the limited data on enteric virus shedding in excretions other than stool. Addressing these gaps will require expanded shedding studies across underrepresented excretions and adherence to standardized reporting framework.

## Contributors

GZ contributed to the conceptualism of the study, data curation, formal analysis, methodology, visualization, writing the original draft, and reviewing subsequent drafts. EMGC contributed to the methodology, data curation, and reviewing subsequent drafts. ABB contributed to the conceptualism of the study, methodology, project administration, visualization, writing the original draft, reviewing subsequent drafts, supervision, and funding acquisition. All authors had full access to all the data in the study and had final responsibility for the decision to submit for publication. All authors verified the underlying data of the study.

## Data sharing

Compiled data are immediately available publicly at the Stanford Digital Repository (https://doi.org/10.25740/cs445xw7641).

