## Appendix for "Systematic review and meta-analysis of enteric virus shedding in human excretions"

### Supplemental methods

**Quality Score.** All papers included in this review were evaluated according to criteria shown in the Data Measurement Quality Assessment Table (Table S2). Each paper was assigned a quality score ranging from 0 to 6, determined by the number of criteria it adhered to. A full point was given to a paper if it fully conformed to a criterion, half a point if it partially met the criterion, and 0 points if it did not adhere at all. If there was uncertainty regarding the paper's conformity to the criteria, it was considered non-compliant and received 0 points. The paper was categorized as low confidence if the paper scored greater than or equal to 0 but less than 2, moderate confidence if it scored greater than or equal to 2 but less than 4, and high confidence if it scored greater than or equal to 4 but less or equal to 6. Data from papers classified as low confidence were excluded from the analysis.

**Assumptions for combining concentration data across studies for each virus and each excretion.** The following assumptions were made. First, samples reported as below the detection limit collected in a cross-sectional study were excluded from this analysis; this is justified because in the cross-sectional study design, patients not infected with the virus may have been included as study participants. Second, for studies reporting summary statistics with no individual-level data, the reported central tendency was used to represent the concentration distribution, weighted by the number of individuals included in the study. If a central tendency was not provided, but a maximum and minimum were, their average was used to represent the central tendency. Third, for patients infected with multiple genotypes of the same virus simultaneously, concentrations of each individual genotype were summed to provide a virus concentration. Fourth, concentration data not reported in externally valid units (virus or viral genome copies per gram or milliliter of excretion), or could not be transformed into externally valid units given information provided by authors, were excluded. Fifth, if necessary for a unit conversation, the density of stool was estimated as 1 g/mL following the US National Bureau of Standards (NBS).[1]

**Assumptions for generating concentration longitudinal profiles.** In addition to the assumptions described in the previous paragraph section, the following assumptions were made. First, concentration data not specified as being collected on a specific day were considered invalid and excluded. Second, for concentration data provided on an hourly time scale, time was categorized such that hours 0 to 23 were designated as Day 0, hours 24 to 47 as Day 1, and this pattern was extended for the following days. Finally, concentration data that fell below the limit of detection were treated as 0 and were consequently included in the analysis.

**Assumptions for generating viral presence longitudinal profiles.** In addition to the assumptions made above for compiling the longitudinal concentration data, the following assumptions were also made. First, for datasets including multiple patients reporting a percentage of positive cases for a specific day, the number of positive specimens for that day was calculated by multiplying the percentage value by the total number of patients in the dataset. Second, when studies reported only summary statistics and did not provide individual-level data, the reported central tendency was used to represent the distribution of virus presence over time, weighted by the number of individuals included in each study.

**Adjusted p value justifications for comparing aggregated virus concentrations.** Two excretion types had concentration data available for two or more viruses (stool and saliva); therefore, we used p=0.025 to account for multiple comparisons. Two viruses had concentration data available for two or more excretions (NoV and HAV); therefore, we used p=0.025.

**Adjusted p value justifications for duration of viral shedding Kruskal Willis tests.** For data categorized by DAS, only two excretions had shedding duration data available for two or more viruses (stool and saliva); therefore, we conducted two Kruskal-Wallis tests and used an adjusted p value of 0.05/2 = 0.025. For data categorized by DAI, only stool has shedding duration data available for two or more viruses, we used a p value of 0.05. Additionally, for each virus, we used the non parametric Kruskal-Wallis test to test the null hypothesis that there is no difference in the shedding duration across excretions. For data sets relative to DAS, two viruses had shedding duration data available for two or more excretions (NoV and RV). Accordingly, two Kruskal-Wallis tests were conducted with an adjusted p value of 0.05/2 = 0.025 to determine significance. For data sets measured with respect to DAI, only NoV has shedding duration data available for two or more excretions, so we used a p value of 0.05.

**Publication bias.** The outcomes in this review (e.g., viral concentration, positivity rates, duration of shedding, and peak day of shedding) do not involve comparisons between two entities (e.g., treatment vs. control groups) or the calculation of effect sizes typically used in intervention studies. Therefore, standard publication bias assessment methods like funnel plots and Egger's regression test are not applicable for this review. To address the potential for publication bias toward selective reporting of positive or negative results, we calculated the proportion of reported positive results as an indicator of potential bias:

$$Proportion\ of\ positive\ results = \frac{\Sigma\ number\ of\ positive\ data\ points\ reported}{\Sigma\ total\ number\ of\ data\ points\ reported}.$$

A proportion between 0 to 0.33 would indicate a selective reporting bias because there is a substantially greater proportion of negative results being reported. A proportion between 0.33 to 0.66 would indicate no publication bias of selective reporting, as studies consistently report both negative and positive results. A proportion between 0.66 to 1 would suggest a bias that indicates null or non-significant findings are not comprehensively reported and there is a risk of overestimating the virus shedding concentration and prevalence of positive outcomes.

### Supplemental results

**Quality Score Assessment.** For NoV, 30 papers were categorized as having high confidence, 39 with moderate confidence, and 1 with low confidence in the measured data. The median quality score was 3 (Figure S1). For RV, 19 papers were deemed to have high confidence, 29 with moderate confidence, and 2 with low confidence in the measured data. The median quality score was 3. For HAV, 12 papers were deemed to have high confidence, 14 with moderate confidence, and 1 with low confidence in the measured data. The median quality score was 3.5. For AdV, 4 papers were classified as high confidence and 7 with moderate confidence in the measured data. The median quality score was 3.25. Papers with low confidence were excluded and not considered in any data analysis.

**Summary of measurement methods and study approaches for NoV.** Out of 66 papers included for NoV, 56 papers utilized PCR, 3 papers utilized ELISA, 2 papers utilized radioimmunoassay, and 5 papers utilized electron microscopy for virus detection and quantification. Among these studies, data were derived from natural NoV infections in 53 papers and human challenge experiments in 13 papers. No data from vaccination studies were included in the analysis.

**Summary of measurement methods and study approaches for RV.** Among the 44 papers included for RV, 24 used PCR, 14 used ELISA, and 6 used electron microscopy for virus detection and quantification. Of these 44 studies, data were derived from natural RV infections (22 papers), human challenge experiments (1 paper), and vaccination studies (21 papers).

**Summary of measurement methods and study approaches for HAV.** Among the 23 papers included for HAV, 14 utilized PCR, 2 papers utilized ELISA, 4 papers utilized radioimmunoassay, and 3 papers utilized electron microscopy for virus detection and quantification. Of these 23 studies, data were derived from natural HAV infections (18 papers), human challenge experiments (2 papers), and vaccination studies (3 papers).

**Summary of measurement methods and study approaches for AdV.** Among the 10 papers included for AdV, 9 papers utilized PCR and 1 utilized electron microscopy for virus detection and quantification. Data from all 10 papers were derived from natural RV infections. No data were derived from either challenge experiments or vaccination studies.

**Summary of studies with paired samples for each virus.** Among the 66 studies included for NoV, 64 reported unpaired data, while 2 presented paired data, where different excretion measurements likely originate from the same patient (Table S7). Similarly, of the 44 studies on RV, 42 reported unpaired data, and 2 included paired data. All studies on HAV and AdV reported only unpaired data. As only 4 papers (less than 3% of the total included studies) contained potentially paired data and omitting them didn't affect the results, we did not differentiate these datasets and included them in the overall analysis.

**Additional details on publication bias.** When examining individual viruses, NoV and RV both had a median proportion of positive results of 0.58 (n = 66 and 44, respectively). For HAV, the median proportion of positive results was 0.51 (n = 23), while for AdV it was 0.41 (n = 10) (Figure S2).

#### Table S1. Search terms and dates used for systematic literature review.

Separate searches were conducted for each virus (n = 4 searches). Each search included the virus search terms and all excretion search terms.

| | Search Item | Search Terms | Date Searched |
|---|---|---|---|
| Virus | Norovirus (NoV) | norovirus* OR norwalk* OR calicivirus* OR "Small Round-Structured Virus*" | 6 Nov 2023 |
| | Rotavirus (RV) | rotavirus* AND human | 11 April 2024 |
| | Hepatitis A virus (HAV) | "Hepatitis A" OR "Hepatitis A virus*" | 29 Nov 2023 |
| | Adenovirus group F (AdV) | “enteric adenovirus*” OR “adenovirus F*” OR “adenovirus 40*” OR “adenovirus 41*” OR "HAdV-F*" OR "HAdV-40" OR "HAdV-41" OR "AdV-F*" OR "AdV-40" OR "AdV-41" | 3 July 2024 |
| Excretion | Stool | feces OR faeces OR stool | |
| | Saliva | saliva | |
| | Vomit | vomit* | |
| | Urine | urine | |
| | Sputum | sputum | |
| | Mucus | mucus | |

**Table S2. Quality Measurement Quality Assessment Criteria.**

| Criteria | |
|---|---|
| 1 | Report sampling methodologies (i.e., where and how samples were collected) |
| 2 | Report sample detection methods (i.e., qPCR or Electron microscopy) |
| 3 | Report detection threshold |
| 4 | Report use of controls (i.e., positive and negative control; half point for only showing one types of control) |
| 5 | Report use of replications |
| 6 | Report sample preservation and processing (i.e., storage and transportation) |

**Table S3. Regression model equations for shedding profile.**

A, B, and C are fitting parameters.

| Regression Model | Model Equation |
|---|---|
| Logistic Model | $log_{10}(Concentration + 1) = \frac{A}{1+e^{B \times (Time - C)}}$<br>$Positivity\ rate = \frac{A}{1+e^{B \times (Time - C)}}$ |
| Gamma Model | $log_{10}(Concentration + 1) = A \times (Time)^{B} \times e^{(-C \times Time)}$<br>$Positivity\ rate = A \times (Time)^{B} \times e^{(-C \times Time)}$ |

**Table S4. Data quality across reviewed studies.**

Abbreviations: NoV = norovirus, RV = rotavirus, HAV = hepatitis A virus, AdV = adenovirus.

| Quality Criteria | NoV | | | RV | | | HAV | | | AdV | | | Overall | | |
|---|---|---|---|---|---|---|---|---|---|---|---|---|---|---|---|
| | Yes (%) | Partial (%) | No (%) | Yes (%) | Partial (%) | No (%) | Yes (%) | Partial (%) | No (%) | Yes (%) | Partial (%) | No (%) | Yes (%) | Partial (%) | No (%) |
| | n (%) | n (%) | n (%) | n (%) | n (%) | n (%) | n (%) | n (%) | n (%) | n (%) | n (%) | n (%) | n (%) | n (%) | n (%) |
| Report sampling methodologies | 48 (72) | 0 (0) | 19 (28) | 35 (76) | 0 (0) | 11 (24) | 18 (75) | 0 (0) | 6 (25) | 7 (70) | 0 (0) | 3 (30) | 108 (73) | 0 (0) | 39 (27) |
| Report sample detection methods | 67 (100) | 0 (0) | 0 (0) | 46 (100) | 0 (0) | 0 (0) | 24 (100) | 0 (0) | 0 (0) | 10 (100) | 0 (0) | 0 (0) | 147 (100) | 0 (0) | 0 (0) |
| Report detection threshold | 48 (72) | 0 (0) | 19 (28) | 22 (48) | 0 (0) | 24 (52) | 14 (58) | 0 (0) | 10 (42) | 5 (50) | 0 (0) | 5 (50) | 89 (61) | 0 (0) | 58 (39) |
| Report use of controls | 17 (25) | 10 (15) | 40 (60) | 12 (26) | 16 (35) | 19 (39) | 7 (29) | 2 (8) | 15 (63) | 4 (40) | 2 (20) | 4 (40) | 40 (27) | 30 (20) | 77 (53) |
| Report use of replications | 18 (27) | 0 (0) | 49 (73) | 11 (24) | 0 (0) | 35 (76) | 3 (13) | 0 (0) | 21 (87) | 1 (10) | 0 (0) | 9 (90) | 33 (22) | 0 (0) | 114 (78) |
| Report sample preservation and processing | 35 (52) | 0 (0) | 32 (48) | 24 (52) | 0 (0) | 22 (48) | 14 (58) | 0 (0) | 10 (42) | 7 (70) | 0 (0) | 4 (40) | 80 (54) | 0 (0) | 67 (46) |

**Table S5. Number of individual data points for each enteric virus and excretion type.**

Abbreviations: NoV = norovirus, RV = rotavirus, HAV = hepatitis A virus, AdV = adenovirus, DAS = days after symptom onset, DAI = days after infection.

| | Stool | Vomit | Saliva |
|---|---|---|---|
| NoV Concentration | 2368 | 104 | 7 |
| NoV Concentration DAS | 438 | 0 | 24 |
| NoV Concentration DAI | 379 | 23 | 0 |
| NoV Presence DAS | 2099 | 8 | 63 |
| NoV Presence DAI | 2397 | 28 | 0 |
| RV Concentration | 988 | 0 | 0 |
| RV Concentration DAS | 4 | 0 | 0 |
| RV Concentration DAI | 362 | 0 | 0 |
| RV Presence DAS | 916 | 0 | 40 |
| RV Presence DAI | 14186 | 0 | 0 |
| HAV Concentration | 104 | 0 | 56 |
| HAV Concentration DAS | 23 | 0 | 0 |
| HAV Concentration DAI | 0 | 0 | 0 |
| HAV Presence DAS | 575 | 0 | 0 |
| HAV Presence DAI | 1576 | 0 | 0 |
| AdV Concentration | 214 | 0 | 0 |
| AdV Concentration DAS | 1 | 0 | 0 |
| AdV Concentration DAI | 0 | 0 | 0 |
| AdV Presence DAS | 12 | 0 | 0 |
| AdV Presence DAI | 0 | 0 | 0 |

#### Table S6. Estimated parameters for logistic and gamma models used to describe viral shedding profiles for longitudinal data.

The table presents parameter estimates (A, B, and C) for different datasets, categorized by virus type, and excretion type. Abbreviations: NoV = norovirus, RV = rotavirus, HAV = hepatitis A virus, AdV = adenovirus, DAS = days after symptom onset, DAI = days after infection, DAD = days after detection.

| | | Parameters | | |
|---|---|---|---|---|
| Dataset | Model | A | B | C |
| NoV - Stool - DAS<br>Longitudinal Concentration | Logistic | 1465 | 0.04055 | -128 |
| NoV - Saliva - DAS<br>Longitudinal Concentration | Logistic | 1.652 | 27.97 | 6.687 |
| NoV - Stool - DAI<br>Longitudinal Concentration | Gamma | 7.90533 | 0.11966 | 0.06331 |
| NoV - Stool - DAD<br>Longitudinal Concentration | Logistic | 3611 | 0.03355 | -183.6 |
| NoV - Stool - DAS<br>Longitudinal Presence | Logistic | 122.74326 | 0.08926 | 12.41957 |
| NoV - Saliva - DAS<br>Longitudinal Presence | Logistic | 81.7972 | 1.3059 | 11.1354 |
| NoV - Stool - DAI<br>Longitudinal Presence | Gamma | 64.39168 | 0.35850 | 0.06576 |
| NoV - Stool - DAD<br>Longitudinal Presence | Logistic | 295.43669 | 0.05366 | -10.90594 |
| RV - Stool - DAI<br>Longitudinal Concentration | Gamma | 5.70998 | 0.28657 | 0.07172 |
| RV - Stool - DAD<br>Longitudinal Concentration | Logistic | 295.43669 | 0.05366 | -10.90594 |
| RV - Stool - DAS<br>Longitudinal Presence | Logistic | 22620 | 0.03793 | -145 |

| | | | | |
|---|---|---|---|---|
| RV - Stool - DAI<br>Longitudinal Presence | Gamma | 38.361702 | 0.481126 | 0.072848 |
| RV - Stool - DAD<br>Longitudinal Presence | Logistic | 10700 | 0.02369 | -204.5 |
| HAV - Stool - DAS<br>Longitudinal Concentration | Logistic | 6.95209 | 0.07015 | 72.85998 |
| HAV - Stool - DAD<br>Longitudinal Concentration | Logistic | 429.5 | 0.01364 | -355.1 |
| HAV - Saliva - DAD<br>Longitudinal Concentration | Logistic | 425.3 | 0.007973 | -620.6 |
| HAV - Stool - DAD<br>Longitudinal Presence | Logistic | 48.7323 | 0.2289 | 94.4389 |
| HAV - Stool - DAS<br>Longitudinal Presence | Logistic | 6792 | 0.01603 | -303.6 |
| HAV - Stool - DAI<br>Longitudinal Presence | Gamma | 4.87328 | 0.42370 | 0.02989 |
| HAV - Saliva - DAD<br>Longitudinal Presence | Logistic | 6247 | 0.006448 | -722 |
| AdV - Stool - DAD<br>Longitudinal Concentration | Logistic | 18.37046 | 0.13308 | 4.61993 |
| AdV - Stool - DAD<br>Longitudinal Presence | Logistic | 103.25104 | 0.20277 | 18.21551 |

**Table S7. Summary of Studies with and without Paired Samples.**

Abbreviations: NoV = norovirus, RV = rotavirus, HAV = hepatitis A virus, AdV = adenovirus, DAS = days after symptom onset, DAI = days after infection, DAD = days after detection.

| | Paper with Paired Samples | Paper without Paired Samples |
|---|---|---|
| Number of Studies for NoV | 2 | 64 |
| Number of Studies for RV | 2 | 42 |
| Number of Studies for HAV | 0 | 24 |
| Number of Studies for AdV | 0 | 10 |

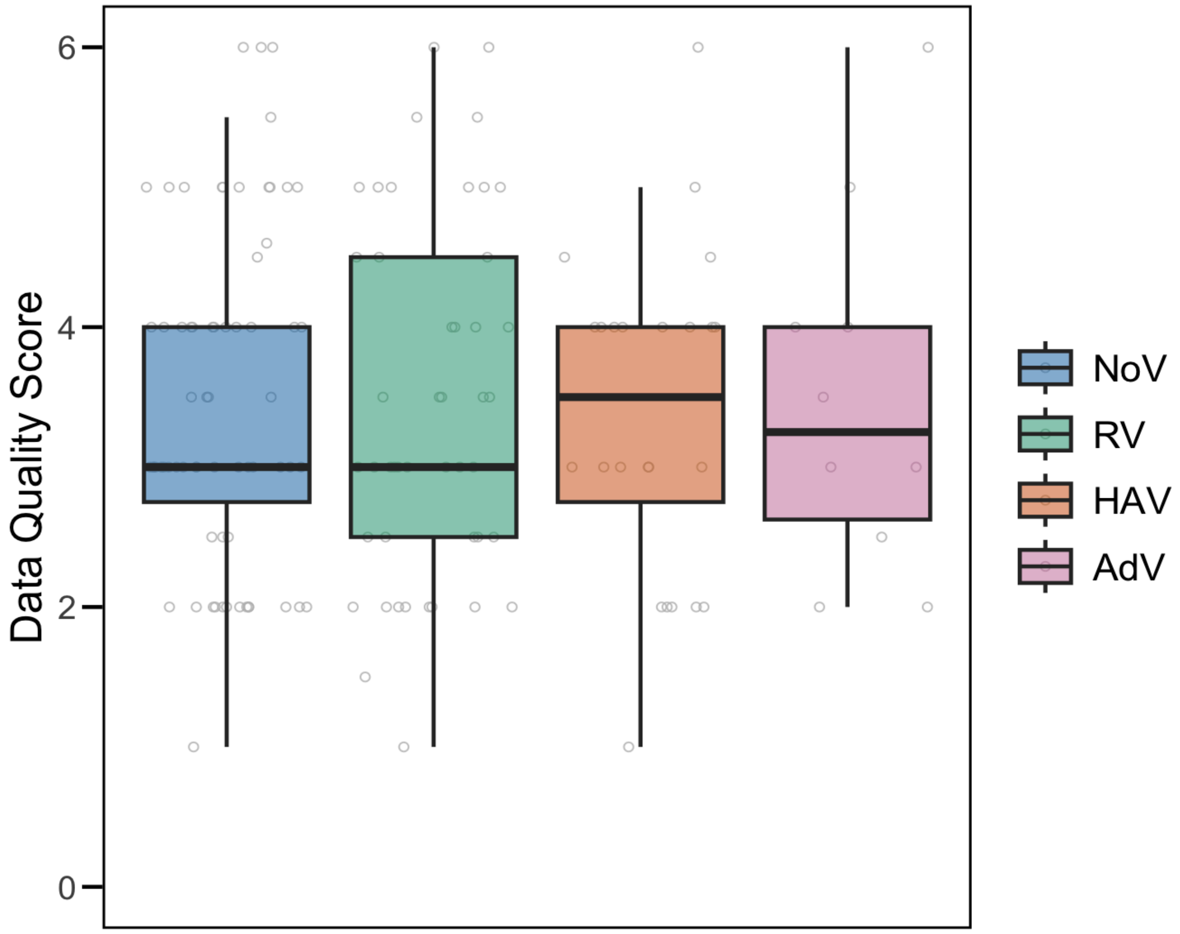

**Figure S1. Data quality assessment score for different enteric viruses.**

Individual data quality score is represented as gray circles. The lower and upper edges of the boxes correspond to the 25th and 75th percentiles, respectively, while the center line represents the median. The upper and lower lines delineate the maximum and minimum, excluding outliers. Abbreviations: NoV = norovirus, RV = rotavirus, HAV = hepatitis A virus, AdV = adenovirus.

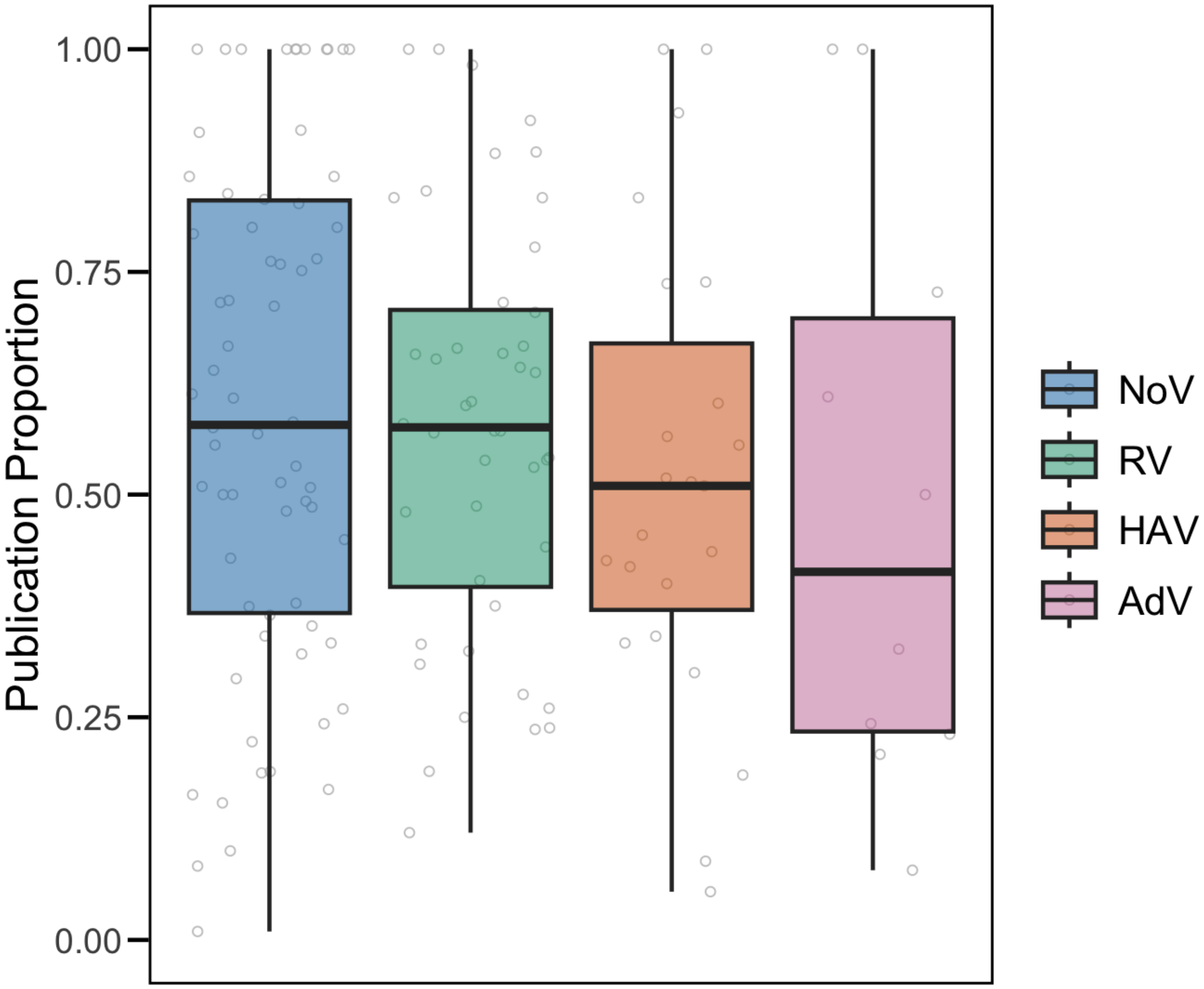

**Figure S2. Publication proportion of positive results for different enteric viruses.**

Individual publication proportion is represented as gray circles. The lower and upper edges of the boxes correspond to the 25th and 75th percentiles, respectively, while the center line represents the median. The upper and lower lines delineate the maximum and minimum, excluding outliers. Abbreviations: NoV = norovirus, RV = rotavirus, HAV = hepatitis A virus, AdV = adenovirus.
